# Association between Lithium and thyroid function: a retrospective study

**DOI:** 10.1101/2024.04.23.24306228

**Authors:** Akanksha Agarwal, Arul Kevin Daniel David, Jayant Mahadevan, Biju Viswanath

## Abstract

**Introduction:** Bipolar Disorder (BD) is associated with thyroid dysfunction, and literature on the clinical impact of Lithium on thyroid function in BD patients remains inconsistent. Thus, we aimed to systematically estimate the prevalence of thyroid dysfunction in Lithium treated BD patients, explore the clinical factors associated with the development of hypothyroidism and time to detection of thyroid dysfunction after Lithium initiation.

**Methods:** A retrospective review of BD patients with a follow-up between Jan to Dec 2019 was conducted. Patients with no pre-existing thyroid dysfunction and at least six months of cumulative exposure to Lithium were included. For the included 444 patients, sociodemographic, clinical variables and laboratory assessment values were charted systematically. Patients with TSH > 5 mIU/L were classified as hypothyroid.

**Results:** 27.7% (n=123) developed thyroid dysfunction; 27.3% (n=121) developed hypothyroidism after Lithium exposure. The median duration of detection of hypothyroidism was 33.6 months for females and 38.4 months for males, with no significant difference across genders (*p=.52)*. A significantly higher proportion of females (49.5%) developed hypothyroidism as opposed to males (23.7%). Hypothyroid females differed significantly with respect to comorbid diabetes mellitus and hypertension; from euthyroid females. First-episode depression was associated with hypothyroidism in males. However, multivariate analyses did not detect any associations.

**Discussion:** One-fourth of Lithium-treated patients developed hypothyroidism. This risk was higher in female patients, with onset close to 33 months, indicating the need for closer monitoring for long durations. The impact of hypothyroidism on prognosis and response to treatment needs further exploration.

## Introduction

Bipolar Disorder (BD) is a chronic disabling psychiatric illness affecting 45 million people worldwide ^1^. The lifetime prevalence of BD in India is 0.5% ^2^. There are clinical differences in BD between Indian and Western samples, with a higher proportion of manic, hypomanic, and mixed episodes than depressive episodes ^3^.

Thyroid dysfunction is associated with BD, with rates higher than those in the general population ^4,5^. The reasons are proposed to be two-fold: a pre-existing higher risk of abnormal thyroid function and the impact of a commonly used mood stabilizer, Lithium. Few studies have also discussed the role of untreated clinical hypothyroidism and, less often, hyperthyroidism on the polarity of mood episodes^5^. A study comparing first-episode BD for thyroid dysfunction noted that 33% of mixed-state patients had elevated TSH levels compared to 7% of patients with mania ^6^, highlighting a possible association between thyroid function and polarity of mood episodes. Most studies have noted the association between the rapid-cycling course of BD and thyroid dysfunction ^7^. It is hypothesized that patients with a pre-existing latent hypothalamic-pituitary-thyroid (HPT) axis dysfunction and thyroid autoimmunity are more likely to experience clinical or subclinical thyroid dysfunction during illness, sometimes due to Lithium, one of the first-line treatments for BD.

Lithium impairs thyroid hormone synthesis and inhibits the release of thyroid hormones from the gland, resulting in increased TSH release via feedback mechanism. It also affects B-cell activity, potentially altering thyroid autoimmunity ^8^. Sometimes the direct toxic effect of Lithium on thyrocytes releases thyroglobulin in the bloodstream ^9^. However, it still remains uncertain whether Lithium plays a causal role in thyroid autoimmunity or simply exacerbates the pre-existing autoimmunity and HPT axis dysfunction.

The literature on the clinical impact of Lithium on thyroid function in BD patients remains inconsistent. A systematic review has stressed that there is no clear association between thyroid autoimmunity and Lithium exposure in BD patients ^7^. Retrospective studies indicate that between 10.3% to 36% of BD patients have elevated TSH values while on Lithium, with variability observed across different populations ^10–12^. Some studies suggest a higher risk in females, the elderly, and individuals with diabetes ^13^. Small case-control studies conducted in India have suggested thyroid gland enlargement and higher average TSH levels in individuals undergoing Lithium treatment ^14,15^. However, prospective studies with follow-up duration between 2-15 years provide inconclusive findings. While some suggest no discernible difference between the general population and those exposed to Lithium, others report an annual incidence of 1.5% in patients undergoing Lithium treatment ^16–18^.

Considering the possible impact of altered thyroid function on illness prognosis, and inconsistencies regarding the effects of Lithium on thyroid function in existing literature, we aimed to systematically estimate the prevalence of thyroid dysfunction in Lithium treated BD patients and explore the clinical factors associated with the development of hypothyroidism. In addition, we examined the time to detection of thyroid dysfunction after Lithium initiation.

### Methodology

We conducted a retrospective review of hospital medical records at the National Institute of Mental Health and Neurosciences (NIMHANS) after approval by the institute ethics committee. As this was a retrospective study and no patient were contacted, written consent was not required.

A list of all the patients diagnosed with BD who had followed up between Jan 2019, and Dec 2019, was accessed from the electronic records.

Records of patients with BD were reviewed if they met the following criteria: a) Diagnosed as BD as per the DSM IV-TR, b) Treatment with Lithium for at least 6 months, irrespective of current treatment with Lithium, c) Thyroid function measured at least once while on Lithium, with either first recorded value being euthyroid or a documented normal TSH before initiation of Lithium. Patients on other drugs affecting thyroid function were excluded. The screening and selection of records were done as explained in Figure 1.

**Figure 1:**
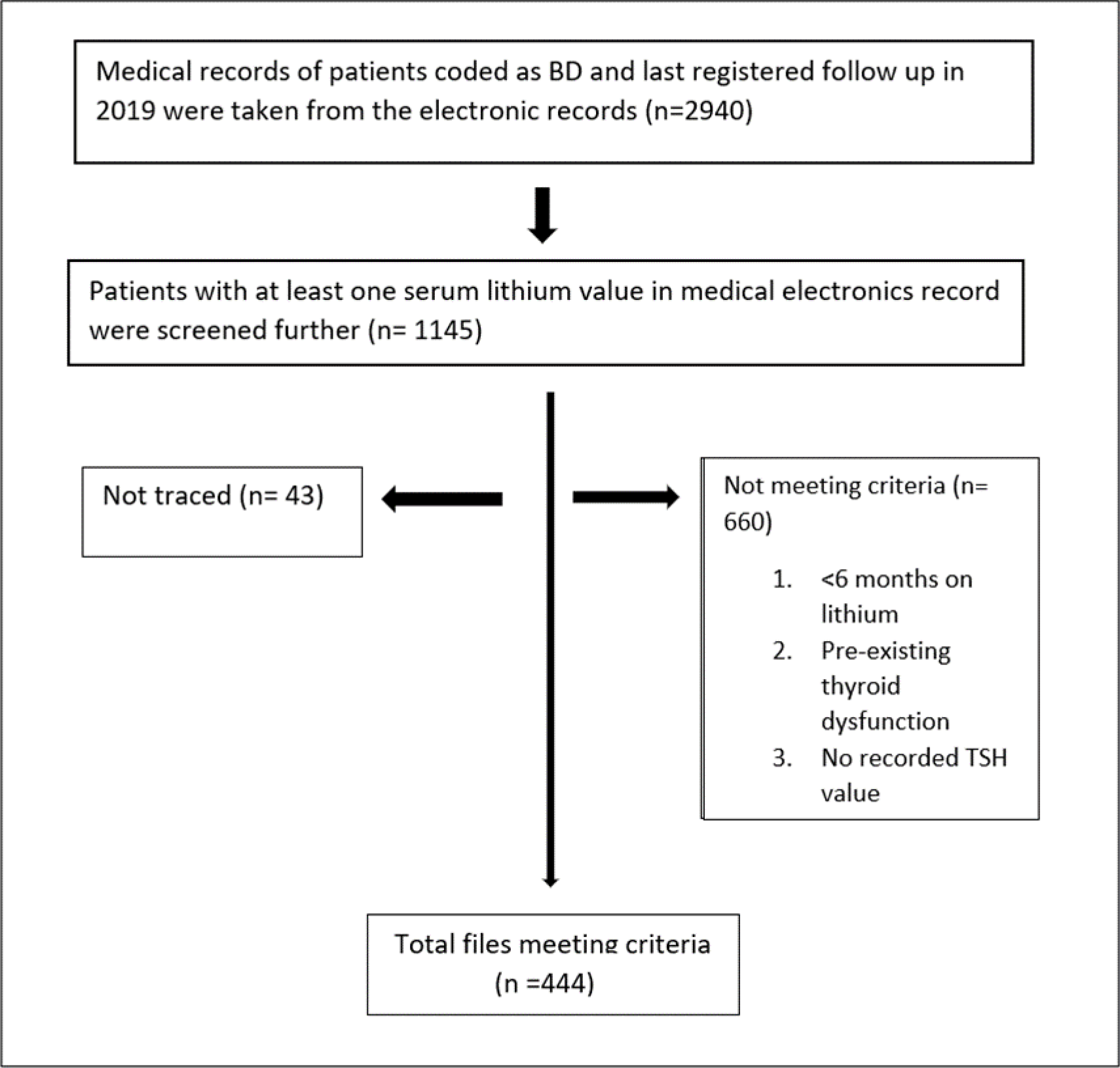
Flowchart depicting the study procedure.

### Clinical profile sheet

The first and second authors reviewed the clinical records of the patients meeting the study criteria (n=444). Sociodemographic and clinical details were obtained from the medical records. National Institute of Mental Health Life chart Method was used to systematically chart the episodes and treatment details ^19^. The clinical variables collected included: age at illness onset, duration of illness, and comorbidities. Treatment details extracted included mood stabilizers, total duration of Lithium use, dose of Lithium, serum Lithium at various doses, and age at initiation of Lithium. The sheet also included details of investigations for thyroid function (T4, T3, TSH) while on Lithium.

### Statistical Analysis

The data entry and analysis were done using SPSS v29 statistical software. Means, medians, and proportions were calculated for continuous variables. The variables were tested for normality using the Shapiro-Wilk test, and the distribution was not normal; hence non-parametric tests were used. Chi-square and Fisher’s exact test analyzed categorical variables wherever appropriate. The results were considered statistically significant if the *p*-value was less than 0.05. A logistic regression was further conducted to determine clinical variables predicting Lithium associated thyroid dysfunction.

## Results

### Sample characteristics

Of 444 patients, 60.8 % (n=270) were males, and 39.2% were females (n=174). The median age of illness onset was 21 years, ranging between 5 and 65 years. The median duration of illness till the last follow-up was 12 years, ranging between 2 years and 44.8 years. The first episode was mania in 68.9% (n=306) and depression in 24.7 % (n=110) patients. For 17.2% (n=30) of women, at least one episode was in the postpartum period. 24.5 % of patients attempted suicide at least once since the onset of illness. 2.3% (n=10) had a rapid-cycling course.

The average duration between the onset of illness and initiation of Lithium was 4.5 years. The median age at initiation of Lithium was 25 years, ranging between 5 to 72 years. The median average serum Lithium level was 0.72 mEq/l.

Ten percent of patients had comorbid diabetes mellitus, 5.4% had hypertension, 2.9% were obese, 10% had comorbid Alcohol use disorder, and 16.3% had Nicotine use disorder.

### Thyroid function characteristics

Thyroid dysfunction was defined as all those who either were started on thyroxine or had a TSH value of >5 m IU/L(hypothyroidism) or < 0.5 m IU/L (hyperthyroidism). Out of 444 individuals, 27.7% (n=123) developed thyroid dysfunction while on Lithium. 27.3% (n=121) developed hypothyroidism while on Lithium. One female patient developed hyperthyroidism after 4 years of Lithium initiation. Euthyroid goiter was noted in 1 female patient after 9 months of Lithium initiation.

The median duration of detection of hypothyroidism since initiation of Lithium was 33.6 months for females, ranging between 1 month to 23 years of cumulative Lithium exposure. For males, the median duration was 38.4 months, ranging between 1 month to 22.5 years. The time to detection of hypothyroidism after Lithium initiation did not differ significantly across genders (*p=.52*).

### Clinical characteristics of euthyroid versus hypothyroid patients

The patients who developed hypothyroidism were compared with euthyroid patients for demographic details, clinical variables, and comorbidities. A significantly higher proportion of females (49.5%) developed hypothyroidism as opposed to males (23.7%); χ2= 4.71, *p*= .03. As sex differences were significant between the hypothyroid and euthyroid groups, further analysis was conducted separately for males and females.

The comparison between euthyroid and hypothyroid males is summarised in Table 1. A significant difference was noted between the groups for the type of first episode. A significantly higher proportion of males with first-episode depression developed hypothyroidism (*p*= 0.008), and there were greater number of depressive episodes in the hypothyroid group (*p*= 0.04).

**Table 1:** Comparison between euthyroid and hypothyroid males.

| Characteristic | Euthyroid | Hypothyroid | Test | *p* |
| --- | --- | --- | --- | --- |
|  | Median (Minimum, Maximum) |  | U |  |
| Age of onset | 21 (8,65) | 22 (5,40) | 6369.5 | .78 |
| Duration of illness(years) | 12.5 (2.4, 43.8) | 12.4 (3.3, 44.2) | 1967.5 | .27 |
| Age at Lithium Initiation | 24 (9,72) | 23.5 (5,45) | 6156.0 | .92 |
| Average serum Lithium | .72 (0.3, 2.6) | .74 (0.3, 1.1) | 6122.5 | .39 |
| | Crosstabulation | | $\chi^2$ | |
| Onset episode |  |  |  |  |
| Depression | 38 | 22 | 7.01 | **.008** |
| Mania | 156 | 39 |  |  |
| Hypertension | 9/206 | 7/64 | 3.78 | .05 |
| Diabetes Mellitus | 20/206 | 7/64 | .081 | .77 |
| Alcohol use disorder | 33/206 | 9/64 | .142 | .71 |
| Nicotine use disorder | 52/206 | 17/64 | .045 | .83 |

The comparison between euthyroid and hypothyroid females is summarised in Table 2. Also, a significantly higher number of females with comorbid diabetes mellitus (p=0.03) and hypertension (p=.017) were detected with hypothyroidism.

**Table 2:**
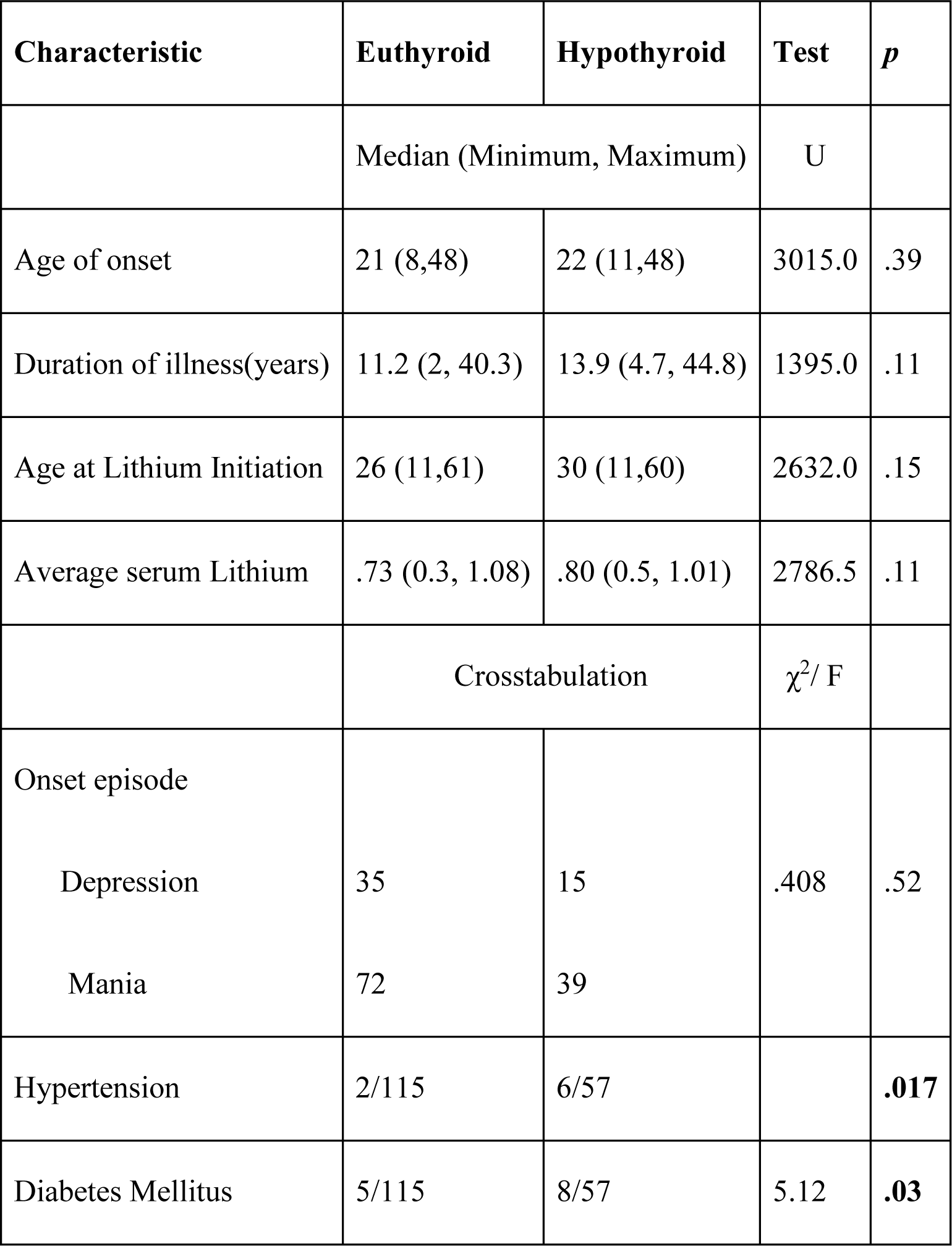
Comparison between euthyroid and hypothyroid females.

| Characteristic | Euthyroid | Hypothyroid | Test | *p* |
| --- | --- | --- | --- | --- |
|  | Median (Minimum, Maximum) |  | U |  |
| Age of onset | 21 (8,48) | 22 (11,48) | 3015.0 | .39 |
| Duration of illness(years) | 11.2 (2, 40.3) | 13.9 (4.7, 44.8) | 1395.0 | .11 |
| Age at Lithium Initiation | 26 (11,61) | 30 (11,60) | 2632.0 | .15 |
| Average serum Lithium | .73 (0.3, 1.08) | .80 (0.5, 1.01) | 2786.5 | .11 |
| | Crosstabulation | | $\chi^2$/ F | |
| Onset episode |  |  |  |  |
| Depression | 35 | 15 | .408 | .52 |
| Mania | 72 | 39 |  |  |
| Hypertension | 2/115 | 6/57 |  | **.017** |
| Diabetes Mellitus | 5/115 | 8/57 | 5.12 | **.03** |

### Clinical factors associated with hypothyroidism

Logistic regression was performed to ascertain the effects of age of onset, age of Lithium initiation, duration of cumulative Lithium exposure, average serum Lithium values, and nature of first episode on the likelihood of detection of hypothyroidism. Table 3 highlights that none of the clinical variables for both males and females predicted the risk of detection of hypothyroidism.

### Clinical predictors of time to detection of hypothyroidism on Lithium

A survival analysis using Cox proportional hazards was conducted for hypothyroid patients to study the role of age of onset, age of Lithium initiation, average serum Lithium, and nature of onset episode on time to detection of hypothyroidism after Lithium initiation. As in Table 4, none of these variables predicted when a patient would be detected with hypothyroidism after initiation of Lithium.

## Discussion

This study systematically examines the prevalence and nature of thyroid dysfunction in BD patients on Lithium. Additionally, we attempt to identify clinical differences between euthyroid and hypothyroid patients.

In our sample, 27.7% of patients developed thyroid dysfunction. 27.3% had hypothyroidism. In the previous studies, the prevalence of hypothyroidism in patients treated with Lithium varied between 0 to 47 % ^5,20^. Geographical and methodological differences across studies can explain the wide variability. These studies also use different criteria for defining clinical and subclinical hypothyroidism. Considering wide variations in the prevalence of hypothyroidism among Indians (11%) ^21^ and the European general population (0.2% to 5.3%) ^22^; our study is instrumental in understanding the additional risk of hypothyroidism due to Lithium exposure in Indian BD patients. Although the prevalence of goiter on Lithium is higher in literature, in our sample, only one patient had goiter^23^.

The median duration of the detection of hypothyroidism on Lithium was 33.6 months for females and 38.4 months for males. months. A retrospective study in the European population has concluded that women were at higher risk of detection of hypothyroidism in the first two years of continuous Lithium exposure ^24^. However, the abnormal TSH in the study was defined as >3.3 mIU/L as opposed to > 5 mIU/L in our study.

We detected a greater incidence of hypothyroidism in female patients, which is clearly supported by existing literature. Although earlier studies reflect higher prevalence in patients with a rapid cycling illness, in our sample, like in other Indian studies ^25^, only 1.8% of patients had a rapid cycling illness. Hence, it is difficult to comment upon its role in thyroid dysfunction.

Overall, there were no clinical factors associated with development of hypothyroidism in the multivariate analysis. However, there were some suggestive findings in univariate analyses which may need further study. A higher proportion of diabetic and hypertensive females developed hypothyroidism. Few of the previous studies describe a higher risk of hypothyroidism in patients with diabetes mellitus ^26–28^ and hypertension ^27,28^. The suggested mechanism for the same is inconclusive; however, it stresses the role of changes in lipid metabolism, genetic factors, and the role of thyroxine in the modulation of insulin resistance. It must be noted that the number of patients with these comorbidities is small to derive clinical significance. Among males treated with Lithium, first-episode depression was associated with the detection of hypothyroidism in males. Males with predominant depressive polarity may have differences in their baseline thyroid autoimmunity, which increases their risk of hypothyroidism when treated with Lithium.

Our study has certain limitations. Some of the patients were treated with other mood stabilizers and atypical antipsychotics. Due to the lack of a control arm, it is difficult to estimate the role of Lithium exposure alone in thyroid dysfunction accurately. Some patients were tested within a few months of initiating Lithium; this duration, however, remains different for all patients. Hence, this reduces the power of the study to detect changes that may have occurred earlier in the course of treatment for some patients. Our sample did not account for a family history of thyroid dysfunction, which independently has a higher risk of thyroid dysfunction. It is possible that thyroid function monitoring was more frequent for patients with higher clinical suspicion of thyroid dysfunction, which could have affected the clinical associations highlighted in the study.

To summarise, one-fourth of Lithium treated patients developed hypothyroidism. This risk was higher in female patients, with onset close to 33 months, indicating the need for closer monitoring for long durations. Whether early detection and treatment of hypothyroidism in these patients impact the prognosis and response to Lithium remains to be studied further.

## Data Availability

All data produced in the present study are available upon reasonable request to the authors.

## Supplementary

**Table 1:**
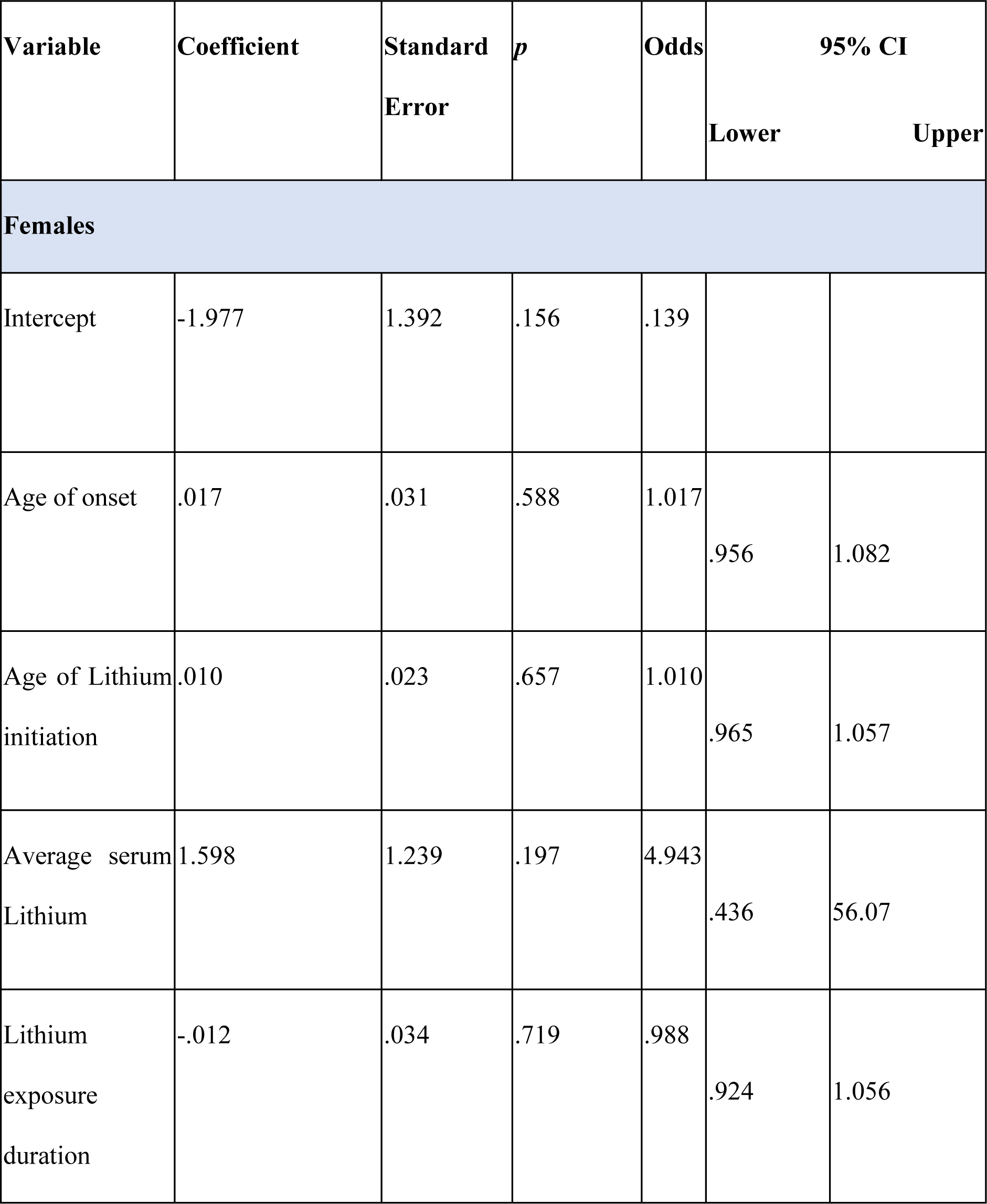
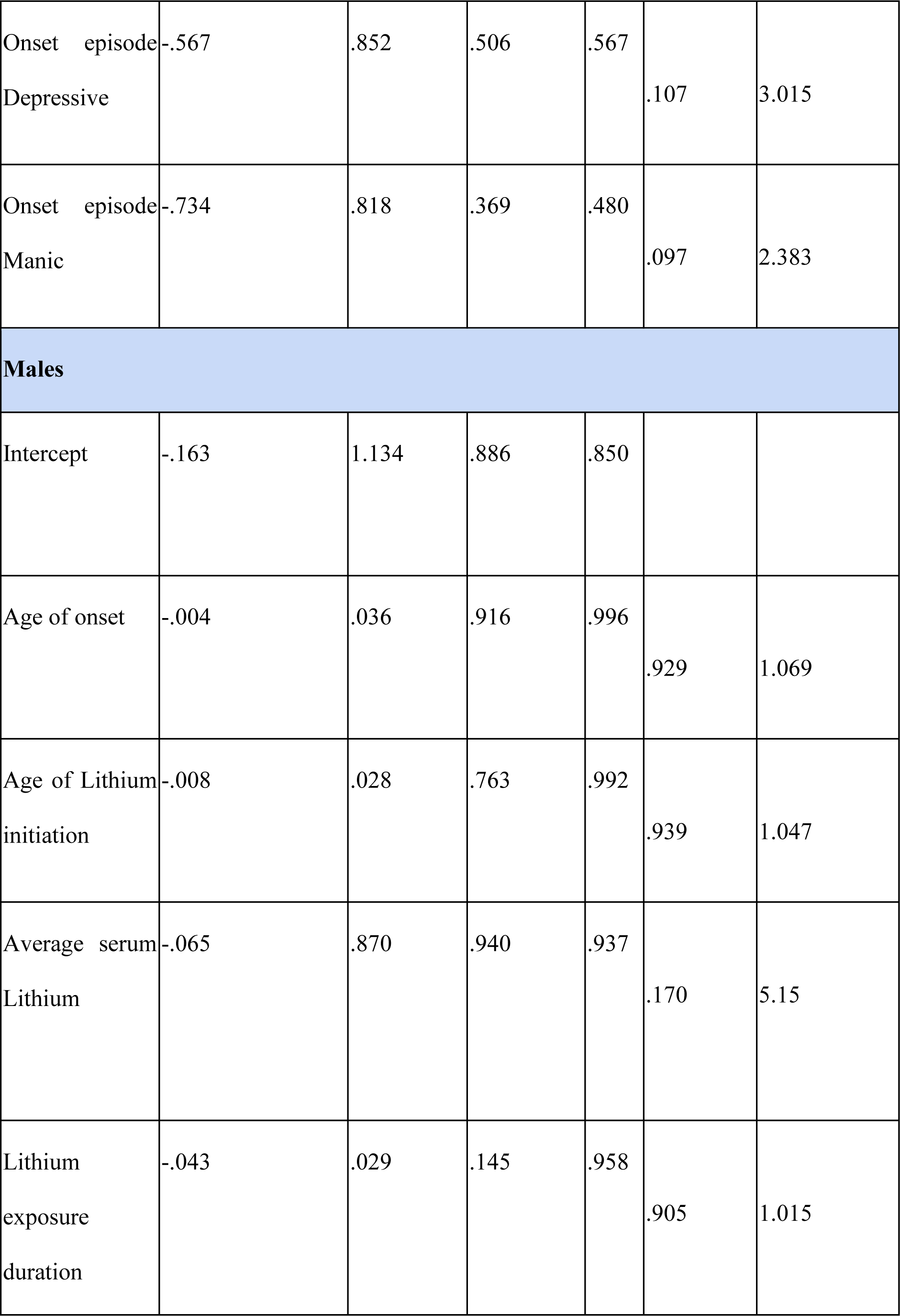
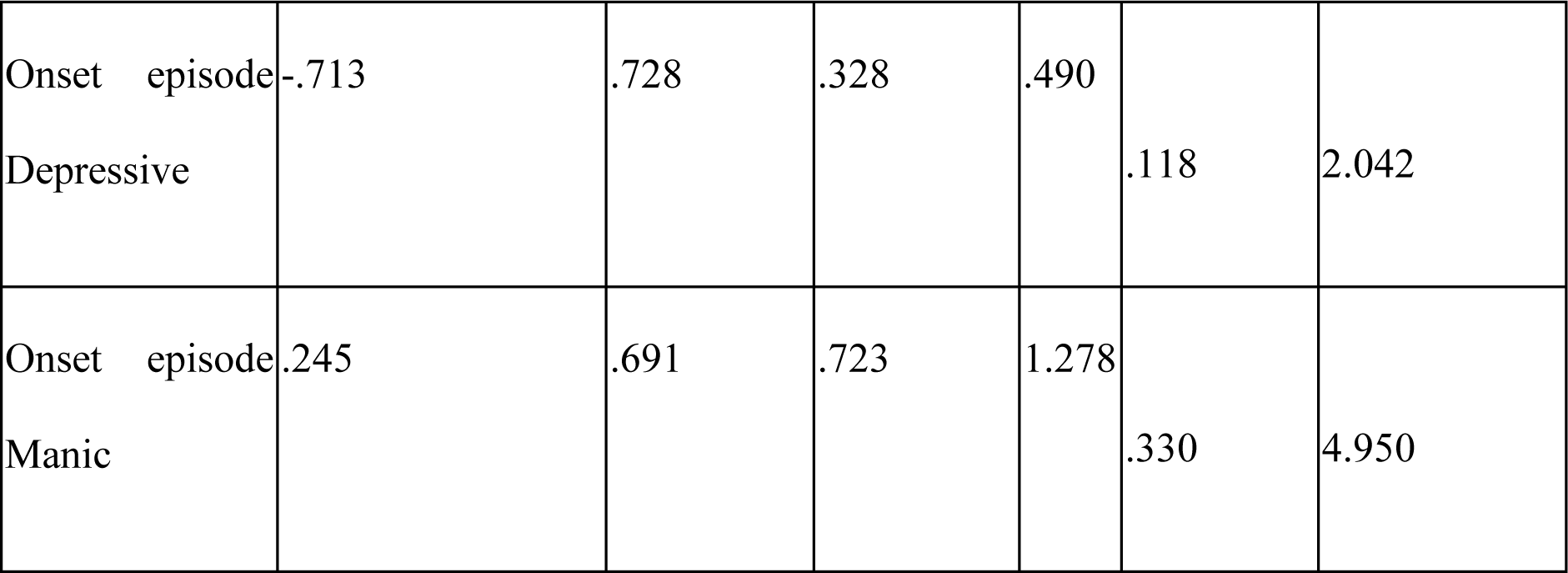
Logistic regression for males and females.

**Table 2:**
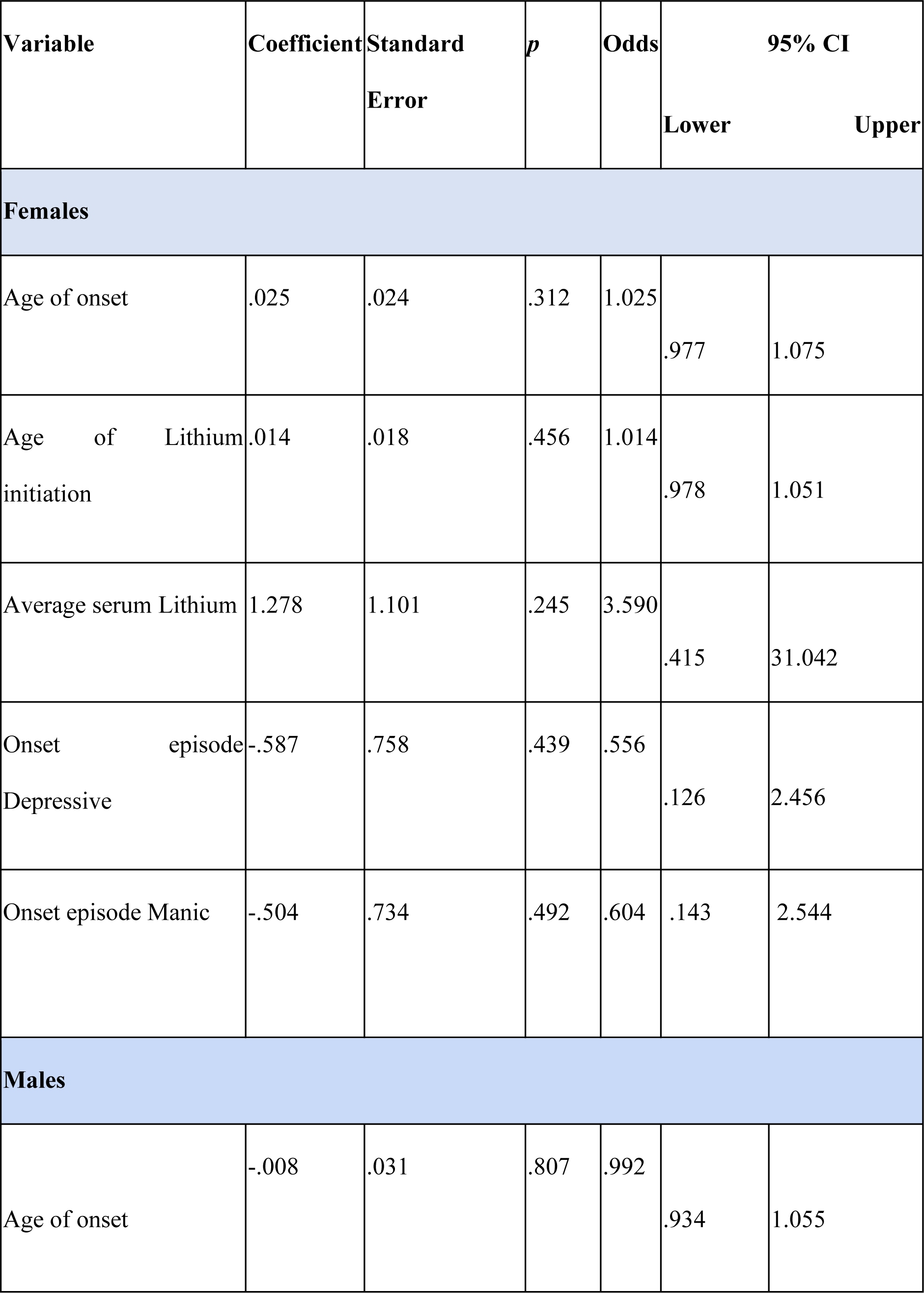
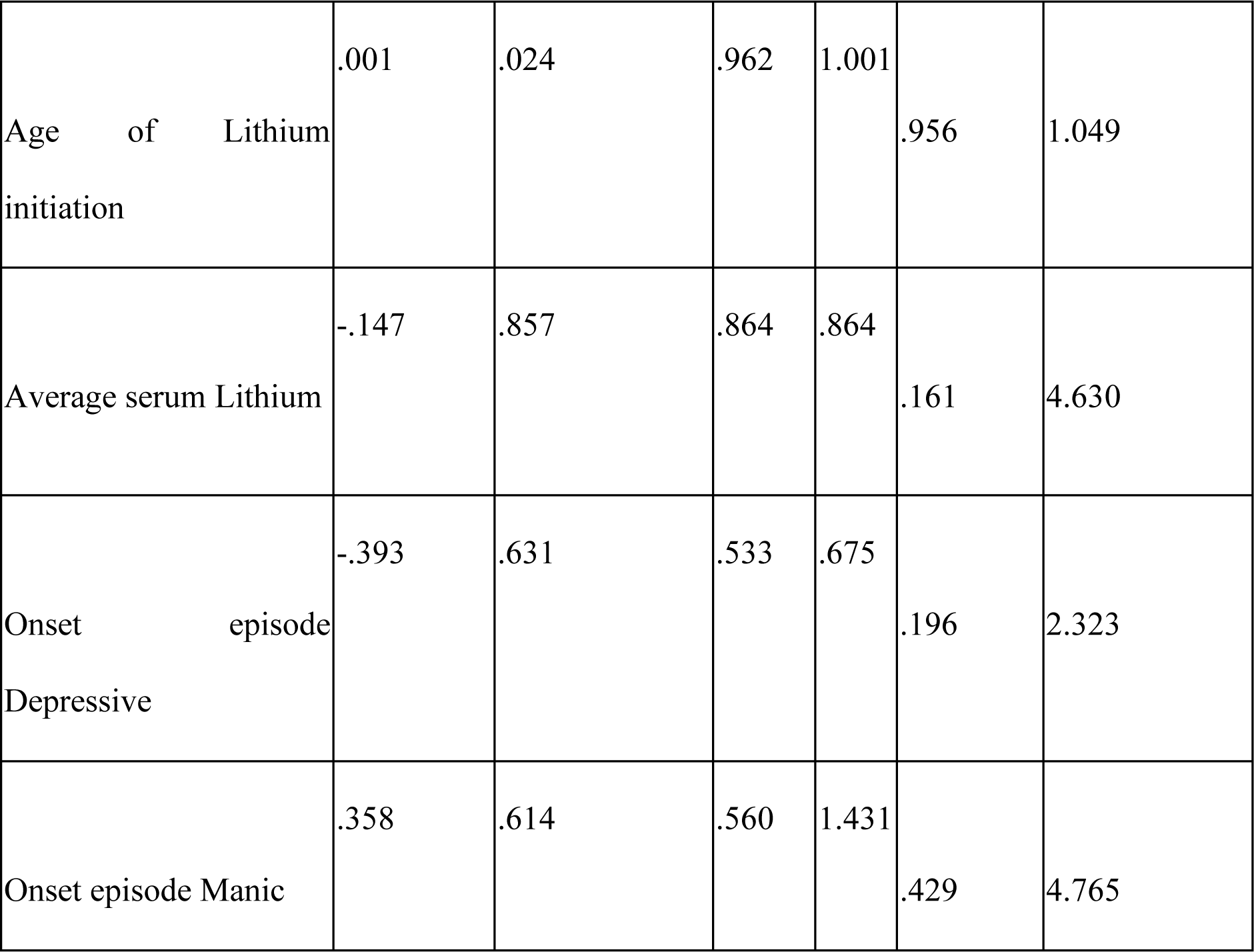
Cox regression for the time to detection of hypothyroidism.

| Variable | Coefficient | Standard Error | *p* | Odds | 95% CI |  |
| --- | --- | --- | --- | --- | --- | --- |
|  |  |  |  |  | Lower | Upper |
| Females |  |  |  |  |  |  |
| Age of onset | .025 | .024 | .312 | 1.025 | .977 | 1.075 |
| Age of Lithium initiation | .014 | .018 | .456 | 1.014 | .978 | 1.051 |
| Average serum Lithium | 1.278 | 1.101 | .245 | 3.590 | .415 | 31.042 |
| Onset episode Depressive | -.587 | .758 | .439 | .556 | .126 | 2.456 |
| Onset episode Manic | -.504 | .734 | .492 | .604 | .143 | 2.544 |
| Males |  |  |  |  |  |  |
| Age of onset | -.008 | .031 | .807 | .992 | .934 | 1.055 |
| Age of Lithium initiation | .001 | .024 | .962 | 1.001 | .956 | 1.049 |
| Average serum Lithium | -.147 | .857 | .864 | .864 | .161 | 4.630 |
| Onset episode Depressive | -.393 | .631 | .533 | .675 | .196 | 2.323 |
| Onset episode Manic | .358 | .614 | .560 | 1.431 | .429 | 4.765 |

